# Deconvolving SARS-CoV-2 mRNA vaccine impact on immunotherapy-related survival in a pandemic

**DOI:** 10.1101/2025.11.21.25340753

**Authors:** Justin Jee, Jiajia Zhang, Jessica A. Lavery, Michele Waters, Christopher Fong, Andy Minn, Michael Glickman, Katherine S. Panageas, Charles L. Sawyers, Nikolaus Schultz

## Abstract

Real-world data suggest that SARS-CoV-2 mRNA vaccines, administered within 100 days of immune checkpoint inhibitor (ICI) treatment (“peri-ICI vaccination”), may improve ICI effectiveness, potentially through synergistic immune priming. Although peri-ICI vaccination was associated with longer survival when we applied a previous framework to our independent dataset, additional patterns emerged. Peri-ICI vaccination benefit diminished after 2021, a pattern confirmed in re-analysis of a published cohort. Benefit extended to patients treated with non-ICI antineoplastics. Benefit also dissipated in landmarked analyses restricted to periods of vaccine eligibility. Finally, progression-free survival in time periods with high vaccine uptake was not longer than in periods without vaccination. These analyses suggest peri-ICI vaccination’s observed association with survival largely reflects selection bias in which patients with better prognosis were more likely to receive SARS-CoV-2 vaccines.

---

A recent study utilizing real-world data has suggested that SARS-CoV-2 mRNA vaccination is associated with improved overall survival (OS) in patients with cancer treated with immune checkpoint inhibitors (ICIs).^1^ Preclinical work further suggested that mRNA vaccination, regardless of tumor neoantigen similarity, induces type I interferon signaling and improves CD8+T cell priming, providing a biologically plausible mechanism by which SARS-CoV-2 vaccination could augment ICI efficacy.^1–3^

However, drawing conclusions, whether associational or causal, from real-world survival data generated during an unprecedented global pandemic is challenging. Disruptions in cancer screening and treatment led to temporary shifts in the characteristics patients receiving therapy.^4^ Population-level behavioral changes, including masking and quarantining,^5^ along with selection bias in those choosing to be vaccinated^6^ collectively increase the potential for biased effect estimates. mRNA vaccines were instrumental in reducing the clinical severity and mortality of COVID-19 in a vulnerable cancer population,^7–9^ presenting an additional potential confounder in any analysis of vaccine effects on cancer mortality in ICI-treated patients.

Here we leveraged a large, deeply annotated cohort^10^ from a tertiary cancer center and, when possible, re-analyzed published datasets, to evaluate the association between SARS-CoV-2 vaccination with survival following cancer treatment initiation. We combined institutionally administered SARS-CoV-2 vaccine and testing records, vaccination data outside our institution reported through health exchanges, clinic visit-associated patient surveys, and AI-assisted natural language processing in free-text clinician notes to maximize ascertainment of vaccination and SARS-CoV-2 infection status. Using this dataset, we first tested the replicability of previously reported findings and confirmed that SARS-CoV-2 vaccination within 100 days of ICI initiation is associated with improved OS when applying an analytic approach as previously published.^1^ We then sought to determine whether:

1. The survival benefit is seen in patients who received SARS-CoV-2 vaccination more than 100 days from the start of treatment.
2. The benefit of SARS-CoV-2 vaccination was seen in later years of the pandemic (post 2021) after vaccination was widespread.
3. The benefit of vaccination was limited to ICI antineoplastics.
4. The survival benefit was seen in landmarked analysis limited only to the period in which all patients were eligible for peri-ICI vaccination.
5. Progression-free survival was improved in a cohort of patients with treatment-naïve stage IV non-small cell lung cancer (NSCLC) during the period when vaccines were available.

## Results

We identified 8,368 patients treated with ICIs between 2017-2022 (characteristics in **Table S1**). A total of 1,410 patients (17%) received SARS-CoV-2 vaccination within 100 days of ICI initiation (“peri-ICI vaccination”). In 2021 alone, 1,588 patients started ICI therapy, of whom 1,377 (87%) had received at least one vaccine dose. This high uptake, consistent with both local^11^ and national^12^ trends, suggests that vaccination status was largely captured in our dataset. As observed in other cohorts,^1^ the number of patients initiating ICI therapy declined in 2020-2021, likely a reflection of pandemic-related disruptions in healthcare delivery. It is noteworthy most patients received COVID-19 vaccination early in 2021, coinciding with the late 2020/early 2021 COVID-19 surge and initial vaccine availability (**Figure S1**).

Applying the previously published analytic specifications^1^ to patients treated from 2017-2022, we likewise observed an association between peri-ICI vaccination and improved OS in multivariable Cox models adjusted for stage, performance status, time since diagnosis, and cancer type, with similar patterns observed within specific cancer types (**Figure S2**). Within the prior analytic framework, we also evaluated the relevance of the 100-day window used to define “peri-vaccination.” Notably, vaccination 100-200 days prior to ICI initiation and vaccination >200 days prior to ICI were similarly associated with longer OS (**Figure S2**), suggesting that the potential association between SARS-CoV-2 vaccination and survival may not be restricted to a narrow time window, as originally proposed.^1^

Cancer treatments, including use of ICIs, have evolved over time, and multiple pandemic-era factors may influence the relationship between vaccination and ICI outcomes. Additionally, the inclusion of 2017-2020 data, a pre-pandemic and primarily pre-vaccine period in which patients initiating ICI were ineligible for peri-ICI vaccination by design, introduces further selection bias.^13^ We therefore examined the association between peri-ICI vaccination and OS by calendar year in 2021 and 2022. Peri-ICI vaccination was associated with longer OS among patients who initiated ICI therapy in 2021 but not among those who started in 2022 (**Figure 1a**, **Figure S3**). To corroborate these findings, we reanalyzed previously published data reporting a survival benefit with peri-ICI vaccination but stratified by year.^1^ In patients with NSCLC treated at MD Anderson, peri-ICI vaccination was likewise not associated with longer OS among those who initiated therapy in 2022, and although the sample size was limited, hazard ratios were close to null (**Figure 1b**). Together, these results suggest that any potential survival advantage associated with peri-ICI vaccination diminished over time, coinciding with a decline in pandemic-era masking and quarantine procedures.^14^

**Figure 1.**
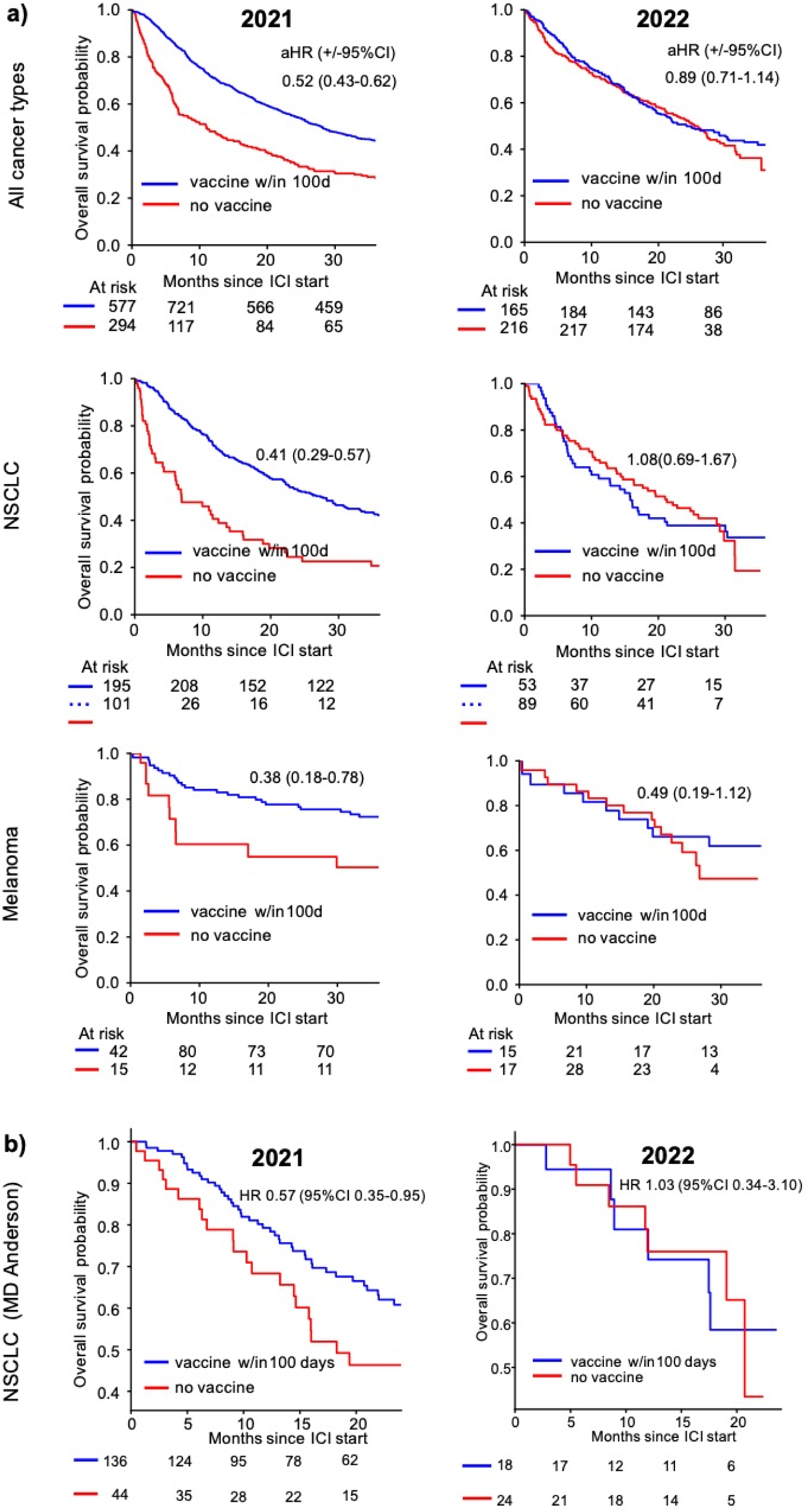
a) Survival following ICI start in 2021 and 2022. Survival is shown by SARS-CoV-2 vaccination status and year of ICI start. Vaccination within 100 days is treated as a time-dependent covariate if given after ICI start. Analyses are risk set adjusted at time of cohort entry (tumor genomic profiling). Adjusted hazard ratios (aHR) for peri-ICI vaccination vs no vaccination are adjusted for cancer type (for “Any cancer type”), stage, time since diagnosis, and performance status. More detailed aHRs by cancer type are presented in **Figure S3**. b) OS following ICI start stratified by peri-ICI vaccination status and year in a previously published cohort of patients with non-small cell lung cancer from MD Anderson. Hazard ratios (HR) are from univariate Cox proportional hazards.

If the benefit of peri-ICI vaccination was driven by synergistic immune priming, the association with improved OS should be limited to patients receiving immune-based therapies. To test this hypothesis, we evaluated OS among patients treated with other commonly administered anticancer agents not co-administered with ICI. Among patients receiving 5-fluorouracil (5-FU), the most frequently used cytotoxic chemotherapy in our dataset, peri-vaccination was associated with longer survival in 2021 but not in 2022, mirroring the temporal pattern observed among ICI treated patients (**Figure 2a**, **Figure S4**). Similarly, we analyzed patients with advanced NSCLC treated with osimertinib, the most common tyrosine kinase inhibitor (TKI) in our dataset. Although the number of patients treated with osimertinib and not vaccinated in 2021 was small, peri-TKI vaccination was associated with longer OS in an analysis combining patients treated from 2017-2021 (**Figure S5**). In 2022, vaccination was no longer associated with benefit (**Figure S5**). These results suggest that the longer OS with SARS-CoV-2 vaccination is not specific to ICI.

**Figure 2.**
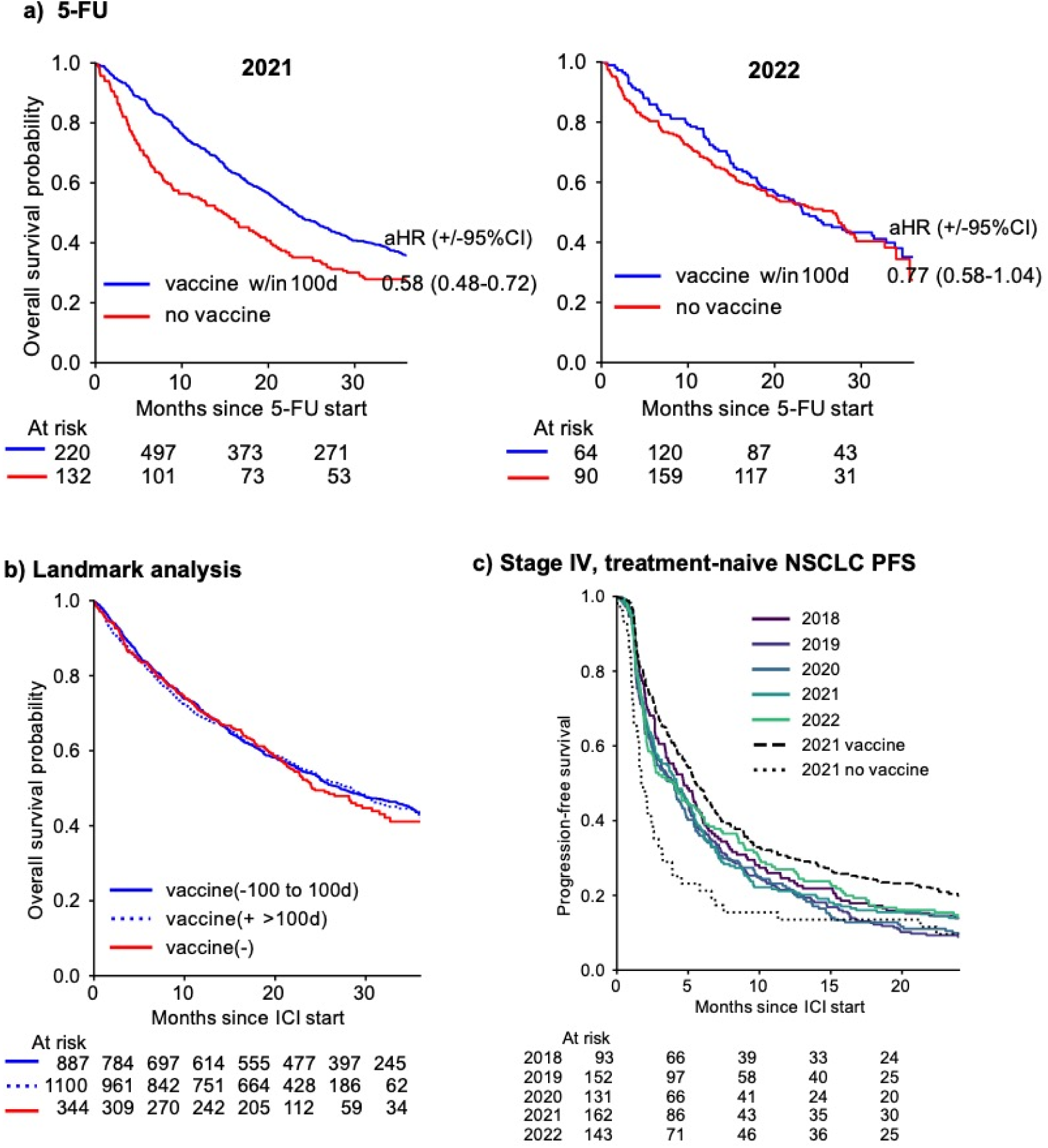
Sensitivity Analyses. a) Overall survival for patients treated with 5-fluorouracil (5-FU) in 2021 and 2022. Vaccination within 100 days is treated as a time-dependent covariate if given after 5-FU start. Analyses are risk set adjusted at time of cohort entry (genomic sequencing). Adjusted hazard ratios (aHR) are adjusted for cancer type, stage, time since diagnosis, and performance status. aHRs for specific cancer types are presented in **Figure S4**. Patients given 5-FU concurrently with immunotherapy were excluded from analysis. b) ICI-treated patients including those treated from March 20, 2025 (i.e. equal eligibility for peri-ICI vaccination) and landmarked at 100 days after ICI start to prevent immortal time bias. c) Progression-free survival (PFS) analysis including only patients with stage IV, treatment naïve NSCLC given ICI within 90 days of diagnosis.

To address several potential biases not accounted for by the previous analytic approach, including a heterogeneous population and changing practices over time in response to the COVID-19 pandemic, we applied analyses landmarked at 100 days after ICI initiation and restricted to March 20, 2021 onward (i.e. 100 days after the vaccine was available). This framework fully captures peri-ICI vaccination and ensures comparable follow-up among vaccinated and unvaccinated individuals. In this analysis, the observed association between vaccination (either peri-ICI vaccination or vaccination >100 days prior to ICI initiation) and survival dissipates (**Figure 2b**). Notably, in 2021 this analysis excludes 21% of patients without peri-ICI vaccination but only 6% of patients with peri-ICI vaccination who died or were lost to follow-up prior to the landmark time. This pattern is consistent with healthier patients being more likely to receive peri-ICI vaccination, especially as it was first available, suggesting that selection bias may contribute to survival differences by vaccination status observed in 2021 in the prior analyses. The potential contribution of immortal time bias was also noted in re-analysis of data from patients with NSCLC from MD Anderson. Although exact treatment start dates were not available to use as inclusion criteria, in this group, peri-ICI vaccination from 2020-2022 was also no longer associated with benefit when analysis was landmarked at 100 days following ICI initiation (**Figure S6**).

If peri-ICI vaccination was beneficial because of a biological immune priming mechanism, then a population with high uptake of SARS-CoV-2 mRNA vaccines should have better outcomes than a baseline population without vaccination. Utilizing OS to test this hypothesis is challenging, as OS may be influenced by use of ICI in earlier stage settings and improving post-ICI lines of therapy over time, such as *KRAS* inhibitors.^15^ To address patient heterogeneity and mitigate the impact of later-line treatments on outcomes, we studied progression-free survival (PFS) in a cohort of patients with treatment-naïve, stage IV NSCLC initiating treatment within 90 days of diagnosis. In this cohort, PFS was similar regardless of year of treatment initiation, including in 2020-2022, inconsistent with the premise that the high uptake of mRNA vaccines would lead to better PFS (**Figure 2c**). Among patients in 2021 treated with ICI, those receiving mRNA vaccines pre- or peri-ICI had better PFS than baseline, while those not receiving mRNA vaccines had worse PFS than patients who initiated ICI before vaccine availability. Together, these results are most consistent with selection bias in which patients with better prognoses were more likely to receive vaccination.

## Discussion

mRNA vaccination has proven a powerful tool for controlling infectious disease^6,16^ and may also hold promise for improving cancer treatment outcomes.^1,17,18^ Preclinical studies offer a compelling biological mechanism by which mRNA vaccination could synergize with ICI treatment in a tumor antigen-agnostic manner. However, estimating the magnitude of SARS-CoV-2 mRNA vaccination effect on long-term cancer outcomes using real-world data remains challenging.

In our analysis, although we replicated the previously reported association between peri-ICI vaccination and improved OS when applying the original analytic approach of a published study,^1^ multiple lines of evidence suggest that this benefit was not specific to ICI and may instead reflect unobserved confounding and selection bias related to the pandemic. The apparent survival advantage extended to vaccinations administered more than 100 days before ICI initiation, timing inconsistent with a mechanistic immune priming mechanism. The benefit of vaccination was absent after 2021 in two independent datasets. Improved survival associated with vaccination in 2021 was also observed among patients receiving non-ICI therapies. Moreover, landmarked analyses restricted to the period in which vaccines were broadly available and PFS analyses in patients with stage IV, treatment-naïve NSCLC did not find a benefit with peri-ICI vaccination. Taken together, these results suggest that vaccination was associated with improved outcomes in a manner specific to the context of the pandemic, likely through selection bias for patients with better prognoses rather than by enhancing anticancer immune responses. Such biases are well-documented in previous vaccination studies^19^ and are likely dependent on practice environment.

A recent randomized trial showed that a tumor antigen-targeting mRNA vaccine, imsapepimut+etimupepimut, showed promise at improving outcomes in patients with melanoma treated with pembrolizumab.^20^ However, the magnitude of benefit from this targeted mRNA vaccine was more modest than that reported in real-world analyses using SARS-CoV-2 vaccination as a presumed immune-priming agent. If mRNA-based immune priming were a universal and robust mechanism, one would expect a neoantigen-directed mRNA vaccine to yield similarly strong, if not greater, improvement in ICI effectiveness.

Limitations of our study include its single center design concentrated in an urban population, a small number of patients treated with certain specific cancer types when stratified by year, and the fact that confounders obscuring relationships between vaccination and ICI outcomes may be unmeasurable. The factors governing immune control of cancer are incompletely understood, and factors like the microbiome^21^ and direct infection exposure^22^ may also incite antitumor immunity. Our data do not eliminate the possibility that in a population under quarantine, with limited microbial exposure, mRNA vaccines may play an outsized role in immune activation, which can in turn affect ICI response. The association between SARS-CoV-2 mRNA vaccines and immunotherapy response may further depend on geographically heterogeneous health practices and shifts in infectious disease exposure before and after COVID-19 lockdowns.^5^ It is also plausible that vaccination altered practice patterns, i.e. delays in diagnosis, procedures, and treatments that stemmed from cautionary measures against COVID-19. These practice delays may have been mitigated in vaccinated patients. We restricted our analysis of peri-ICI vaccination to patients treated through 2022 because declining vaccination rates from 2023 onward are difficult to interpret as reflecting national trends or incomplete data capture. Lastly, analyses do not account for subsequent treatments received, which may have differed among vaccinated and unvaccinated patients, and may contribute to estimates of overall survival in both groups.

Promising preclinical evidence suggests that antigen-agnostic mRNA-based innate immune stimulation may improve ICI effectiveness.^1^ These findings align with recent studies demonstrating the value of immune system priming such as the use of Bacillus Calmette-Guérin vaccines in bladder cancer^23^ and TLR 7/8 agonists in NSCLC.^24^ However, our results indicate that pandemic era associations of peri-ICI vaccination with ICI survival may be attributable to selection bias, and in the post-pandemic era, peri-ICI vaccination is not associated with longer OS. Therefore, speculation based on recently reported real-world outcome data that antigen-agnostic mRNA vaccines may be as or more promising than antigen-tailored approaches should be reconsidered. More broadly, these findings highlight the need for caution when interpreting observational data generated during an unprecedented pandemic.

## Materials and Methods

### Patients

We studied patients treated at Memorial Sloan Kettering Cancer Center (MSK), a tertiary cancer center in New York, NY. The study included patients in the MSK Clinicogenomic Harmonized Oncologic Real-world Dataset (MSK-CHORD) who initiated treatment with ICI, 5-FU, or Osimertinib (NSCLC only) between Jan 1, 2017 and Dec 31, 2022.^10^ Vital status was last ascertained on Feb 1, 2025. To reduce confounding by use of treatments in atypical cancer types, for immune checkpoint inhibitor (ICI) and 5-fluorouracil analysis, only cancer types with at least 500 patients treated with that respective medication were included. Patients receiving concurrent 5-FU and ICI were only included in the ICI analyses. The study was approved by the MSK Institutional Review Board, which waived the requirement for informed consent given the retrospective nature of the analysis.

### Data Annotation

Treatment, performance status, radiographic progression, diagnosis time, stage, and cancer type were annotated using previously described methods.^10^ For progression-free survival analyses, an event was defined as the first radiographic progression ≥30 days after ICI initiation or death, whichever was first. Patients were right-censored at date of last follow-up. SARS-CoV-2 vaccination data including dates were obtained from a combination of structured survey, institutional testing and administration records, health exchange data from vaccination data outside MSK, and free-text reports analyzed by AI large language models (LLMs, Claude Sonnet 4.5). Clinician review (by JZ and JJ) of 20 randomly selected vaccination cases found two cases in which vaccination status may be a false negative and 20 negative cases, none of which were deemed positive by the model. In both false negative cases, there was disparate documentation across an individual patient’s health record regarding vaccination status. All dates in the manually reviewed subset were abstracted correctly by the LLM.

### Statistical Analysis

Peri-ICI vaccination status was defined as receipt of a SARS-CoV-2 mRNA vaccination within 100 days before or after treatment initiation. Overall survival by vaccination status was estimated using Kaplan-Meier methods and Cox proportional hazards models. Patients alive at the end of the study period were censored at their last visit date. Because tumor genomic profiling with MSK-IMPACT was an inclusion criterion for MSK-CHORD, all analyses were risk-set adjusted at time of first tumor sequencing.^25^ Multivariable Cox proportional hazards were fitted including vaccination status, disease stage, performance (ECOG, as a continuous variable), and time since diagnosis across cancers and in specific cancer subtypes. Pan-cancer analyses were also adjusted for cancer type. Fewer than 8% of patients in each treatment cohort had missing data in at least one variable (either time since diagnosis or performance status) and were excluded from multivariable analysis.

When replicating previously published analyses, overall survival was estimated from treatment initiation and peri-ICI vaccination was treated as a time-dependent covariate for patients receiving peri-ICI vaccination after ICI start (patients vaccinated more than 100 days after treatment initiation remained in the no peri-ICI group). Patients vaccinated within the 100 days prior to treatment initiation were in the peri-ICI group at baseline. To address potential immortal time and selection bias related to peri-ICI vaccine administration, analyses were also performed among patients whose treatment started March 20, 2021 onward using a 100-day landmark after treatment initiation. Patients who died or were lost to follow-up prior to 100 days were excluded from these analyses.

Secondary analyses using the initial framework were performed defining vaccination status as (1) peri-ICI SARS-CoV-2 vaccination (2) previous vaccination up to 200 days prior to ICI initiation, and (3) no previous or peri-ICI vaccination. For patients with multiple vaccinations within the window, the vaccination closest to treatment initiation was selected. Analyses were performed in Python 3.10.11 using lifelines 0.26.0.

## Data Availability

Data will be made available upon formal publication of the manuscript

## Acknowledgment

We acknowledge useful discussions with Marc Ladanyi as well as members of the Jee and Schultz labs.

---

**Figure S1.**
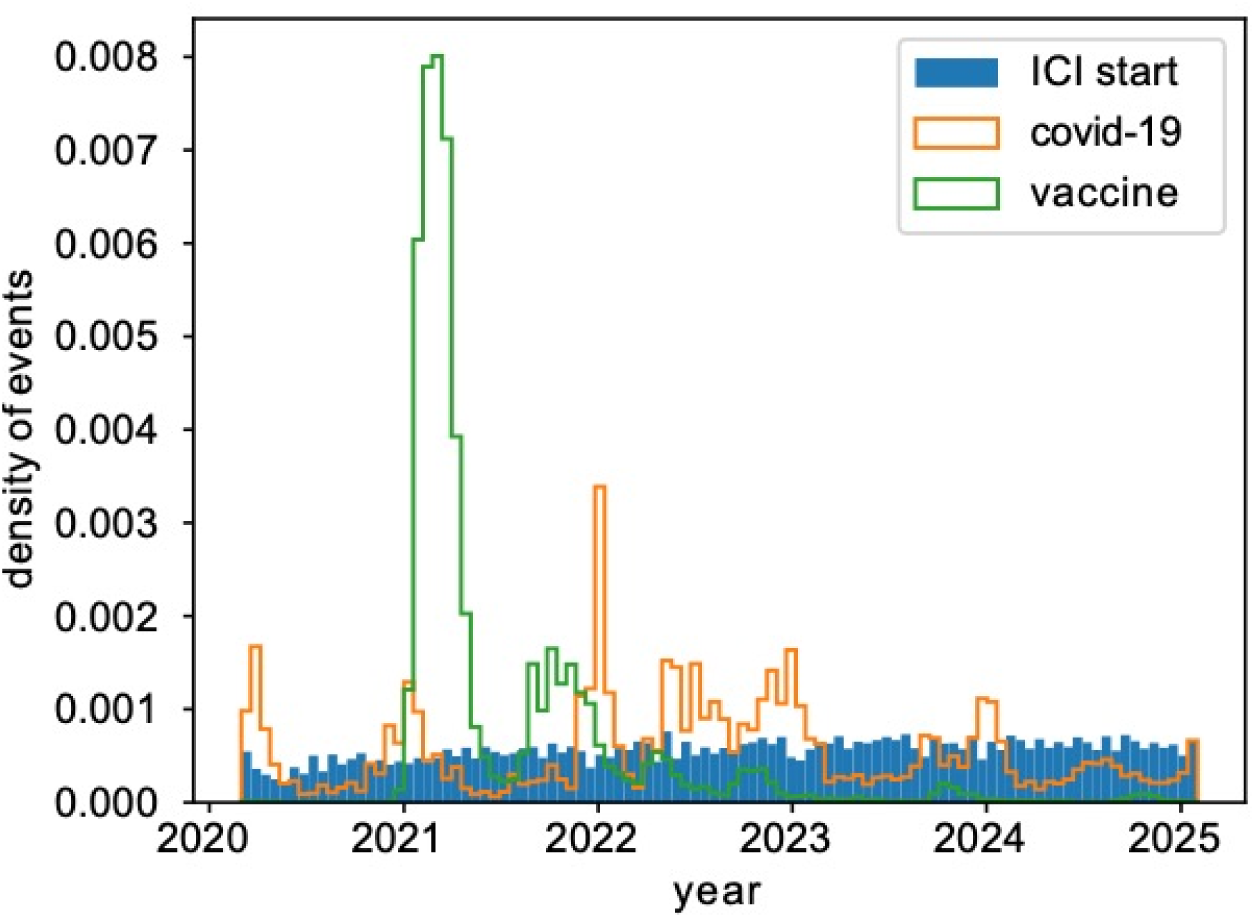
Changing event rates over time. SARS-CoV-2 infection and vaccination rates, as well as immune checkpoint inhibitor (ICI) administration over time

**Figure S2.**
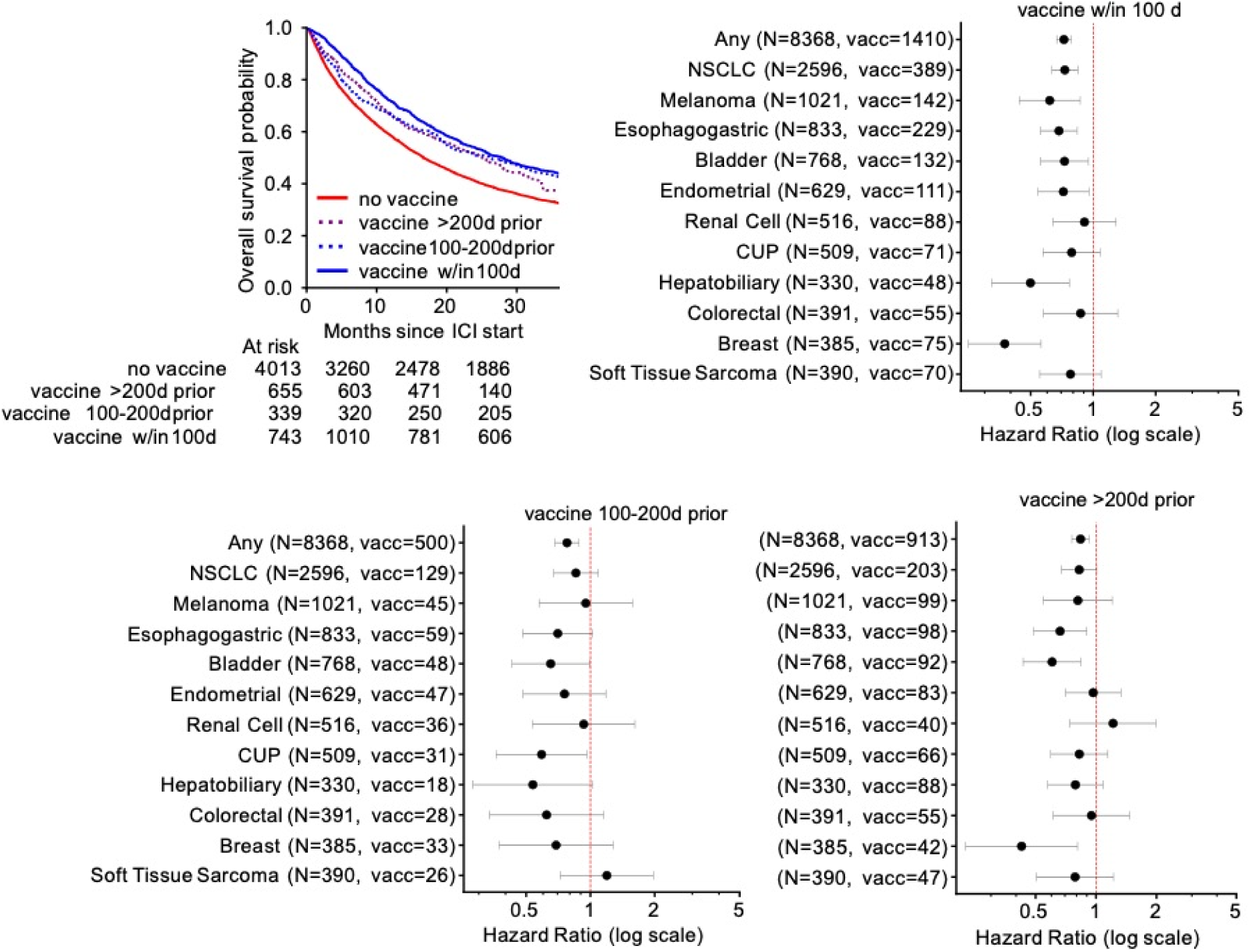
Survival following ICI start stratified by SARS-CoV-2 vaccination status from 2017-2022. Kaplan-Meier overall survival curve for all included patients is shown, with peri-ICI vaccination events after ICI start treated as time-dependent covariates and risk set adjusted at time of cohort entry (tumor genomic profiling). Forest plots show adjusted Cox proportional hazard ratios +/− 95%CI for peri-ICI vaccination, 100-200 days before ICI start, >200 days before ICI start, and without peri-ICI vaccination stratified by cancer type. Hazard ratios are adjusted for disease stage, ECOG, and time since diagnosis.

**Figure S3.**
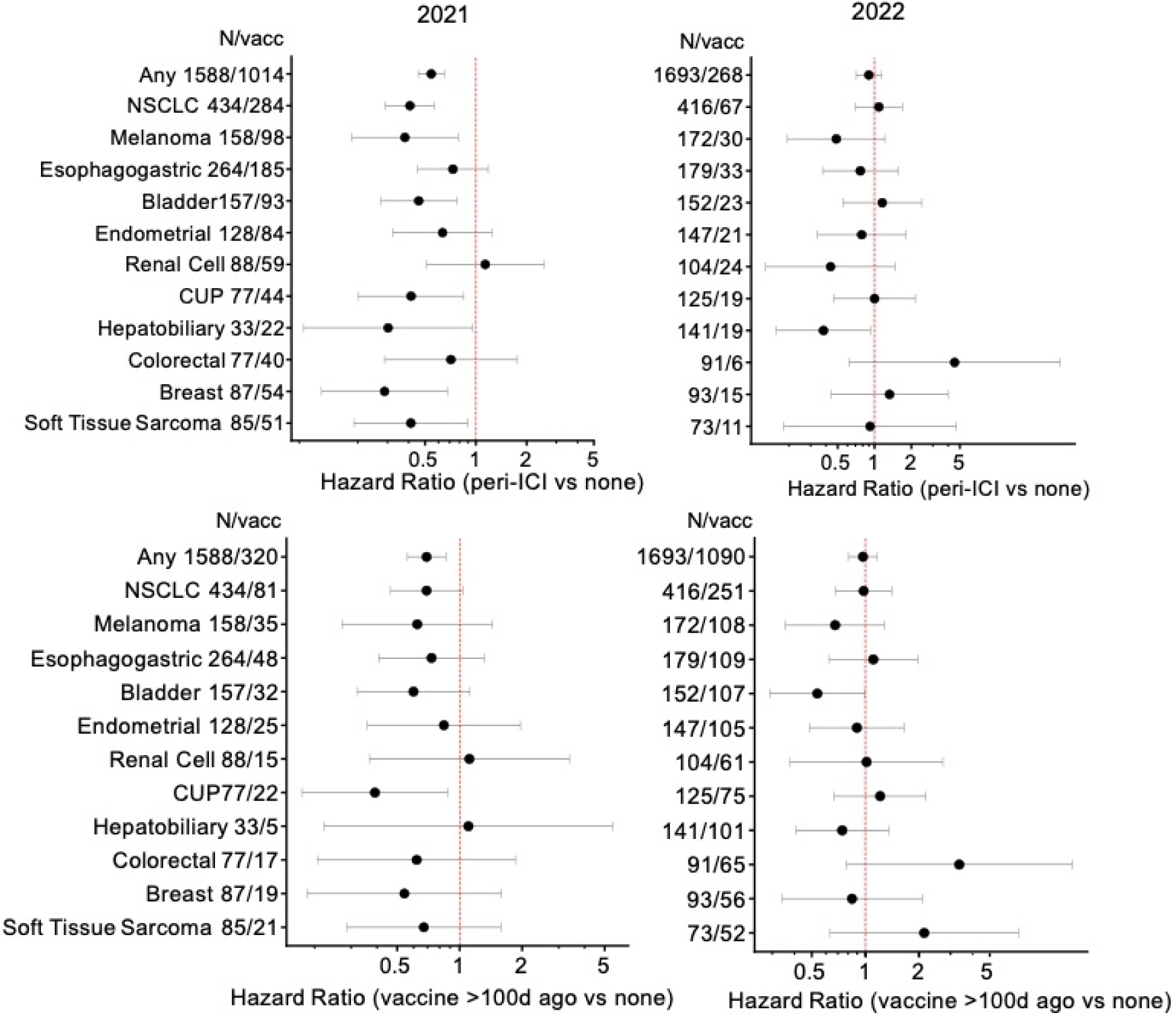
Forest plots show hazard ratios for peri-ICI vaccination and are adjusted for disease stage, ECOG, prior vaccination, and time since diagnosis. “Any” hazard ratio is also adjusted for cancer type. NSCLC=non-small cell lung cancer. CUP=cancer of unknown primary.

**Figure S4.**
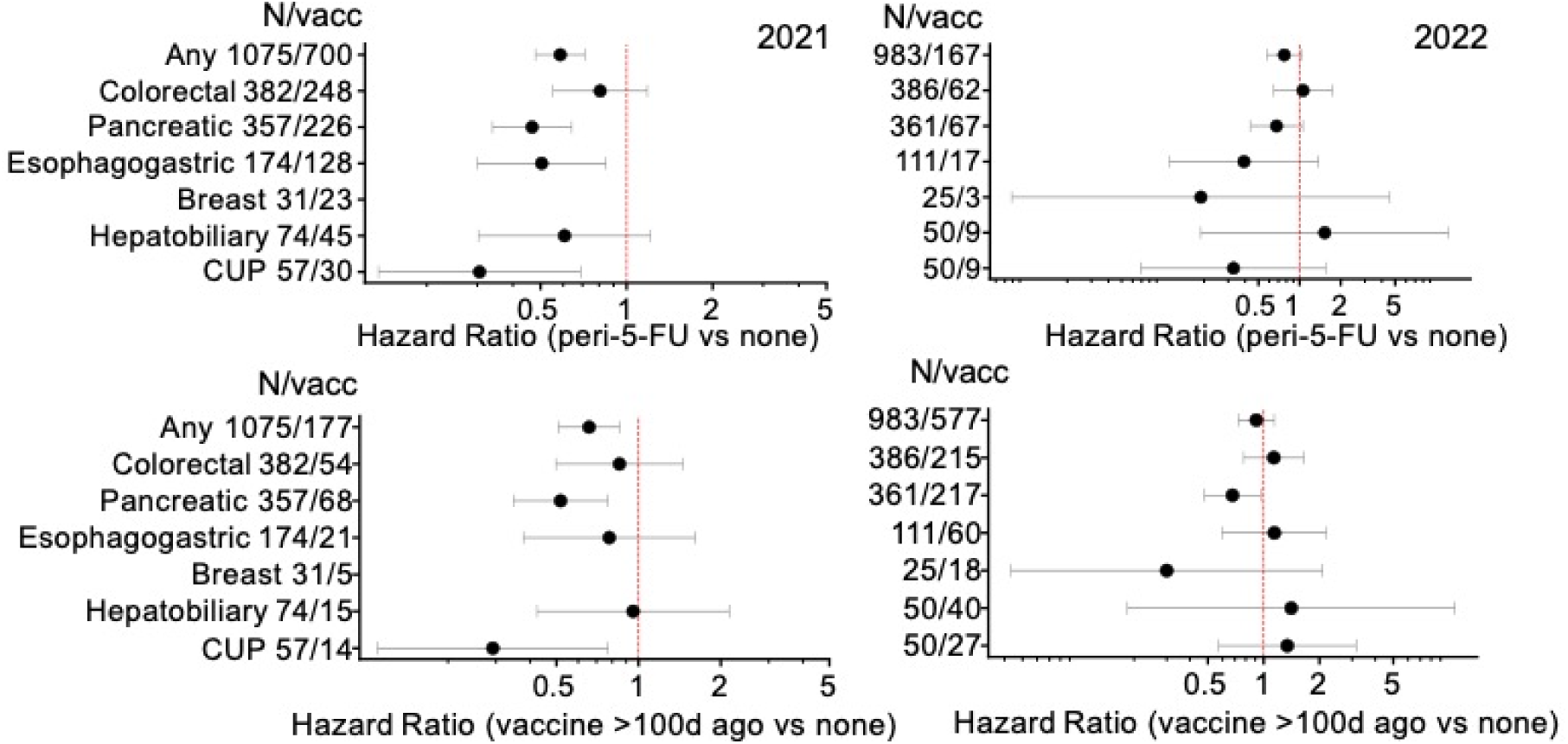
Forest plots show hazard ratios for peri-5-FU vaccination and are adjusted for disease stage, ECOG, prior vaccination, and time since diagnosis. “Any” hazard ratio is also adjusted for cancer type. CUP=cancer of unknown primary. Patients treated with simultaneous ICI are excluded.

**Figure S5.**
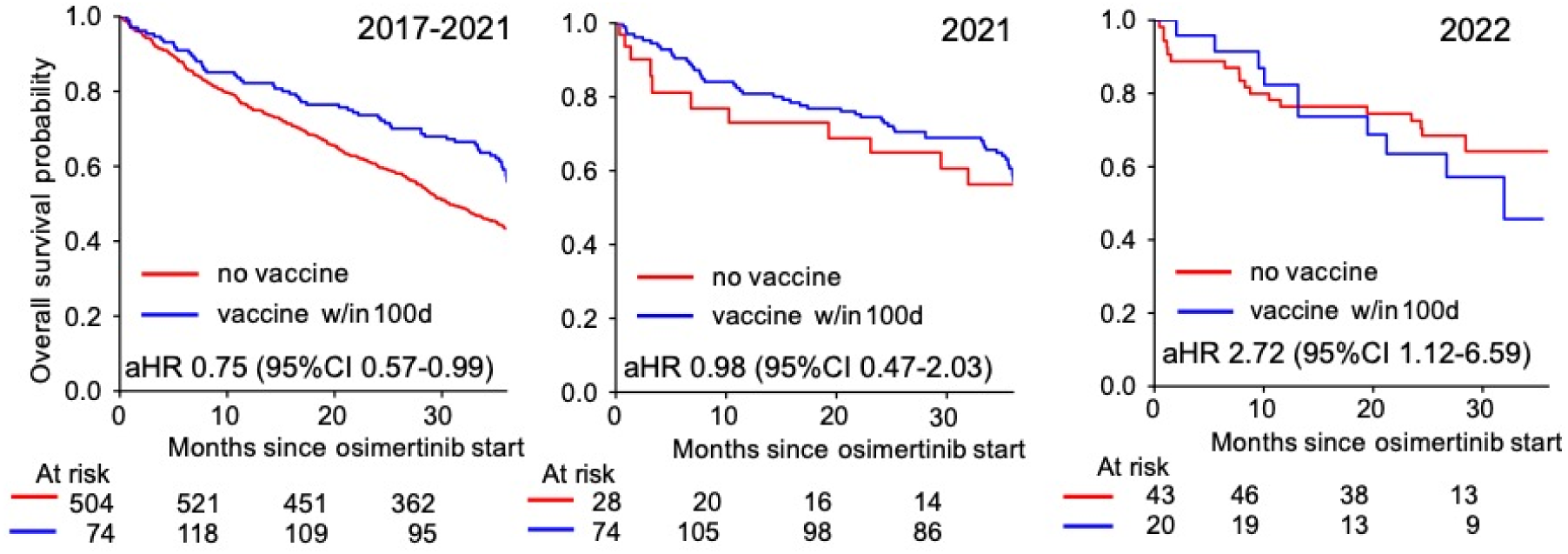
Overall survival for patients treated with osimertinib. Data is stratified by the years shown. Vaccine within 100 days is treated as a time-dependent covariate if given after treatment start. Analyses are risk-set adjusted at time of cohort entry (tumor genomic profiling). Hazard ratios are adjusted for stage, time since diagnosis, and performance status.

**Figure S6.**
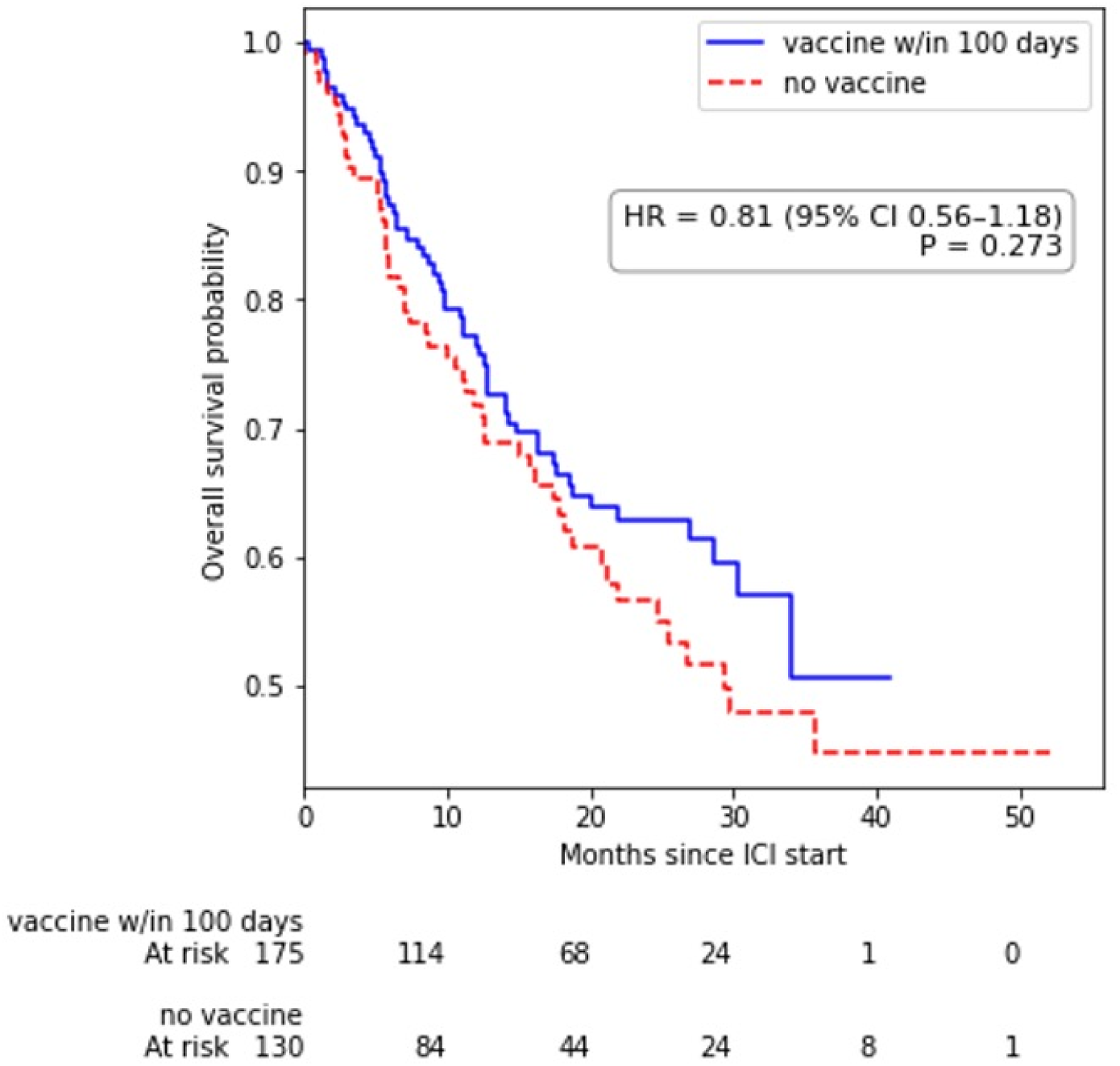
Landmarked analysis in the MD Anderson cohort (NSCLC) starting ICI 2020-2022. Landmarking is done at 100 days following ICI initiation to prevent immortal time bias.

**Table S1.**
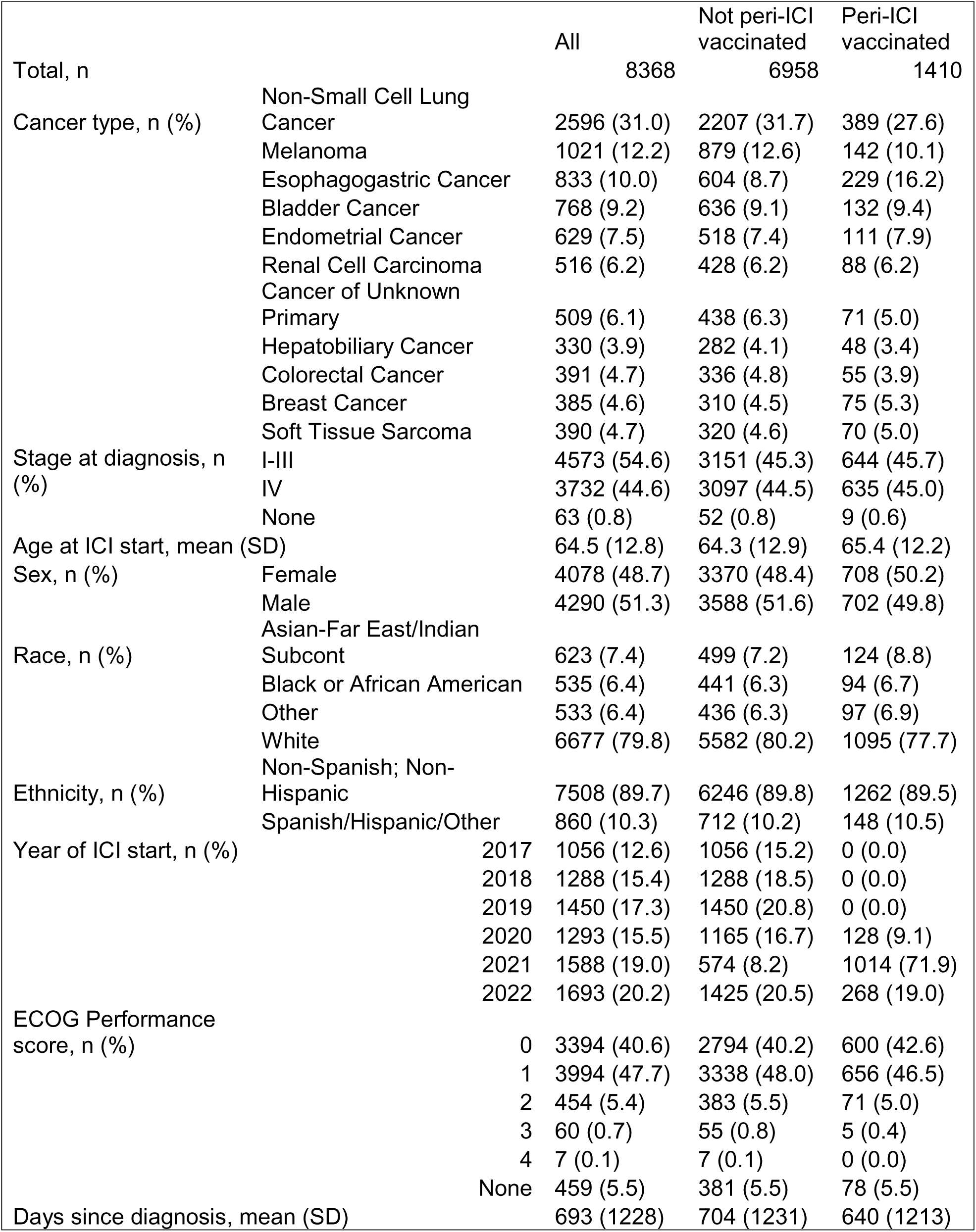
Characteristics of patients treated with immune checkpoint inhibitors (ICI). N (column-wise category %)

